# Environmental risk factors of airborne viral transmission: Humidity, Influenza and SARS-CoV-2 in the Netherlands

**DOI:** 10.1101/2020.08.18.20177444

**Authors:** Edsard Ravelli, Rolando Gonzales Martinez

## Abstract

**Objective:** The relationship between specific humidity and influenza/SARS-CoV-2 in the Netherlands is evaluated over time and at regional level.

**Design:** Parametric and non-parametric correlation coefficients are calculated to quantify the relationship between humidity and influenza, using five years of weekly data. Bayesian spatio-temporal models—with a Poisson and a Gaussian likelihood—are estimated to find the relationship between regional humidity and the daily cases of SARS-CoV-2 in the municipalities and provinces of the Netherlands.

**Results:** An inverse (negative) relationship is observed between specific humidity and the incidence of influenza between 2015 and 2019. The space-time analysis indicates that an increase of specific humidity of one gram of water vapor per kilogram of air (1 g/kg) is related to a reduction of approximately 5% in the risk of COVID-19 infections.

**Conclusions:** The increase in humidity during the outbreak of the SARS-CoV-2 in the Netherlands helped to reduce the risk of regional COVID-19 infections. Public policies that promote higher levels of specific humidification—above 6 g/Kg—can lead to significant reductions in the spread of respiratory viruses, such as influenza and SARS-CoV-2.

## 1. Introduction

Previous experimental evidence indicates that that the aerosolization of secretions lubricating the vocal cords can be a major source of microscopic droplet (microdroplet) virus infections [1]. Evidence of airborne spread of the severe acute respiratory syndrome virus (SARS) is provided by [2]. Recent studies also suggest the possibility of airborne transmission of SARS-CoV-2 through respiratory air droplets and aerosols [3].

Weather factors—such as humidity and temperature—are suspected of playing a role in the transmission of viral particles through aerosolized droplet nuclei or aerosols. Particularly, differences in humidity are suspected to provide an explanation for the observed variability of influenza and its transmission [4]. Weather conditions can have a similar effect on SARS-CoV-2 (COVID-19), as airborne inactivation of the human coronavirus 229E is affected by temperature and relative humidity [5], and in experimental settings both temperature and absolute humidity affect the environmental survival of surrogates of mammalian coronaviruses [6].

Based on observational data, the relationship between SARS-CoV-2 and weather has been recently analyzed in published studies [7-12] and pre-prints [13-18]. All of these studies find a negative correlation between absolute humidity and the spread of SARS-CoV-2, indicating that low levels of humidity increase the risk of COVID-19 cases. In [18], for example, weather conditions are investigated worldwide, and [18] conclude that the community transmission of SARS-CoV-2 is consistent with average temperatures of 5° to 11°C, combined with low specific humidity between 3 to 6 g/kg. Humidity seems to be the most important factor in viral spreading, while temperature play a less important role [13].

Our study provides new evidence about the relationship between specific humidity and the risk of influenza and COVID-19 cases. Five years of hourly weather data from 34 regional weather stations in the Netherlands is analyzed against five years of weekly data of influenza cases, as well as daily regional data of COVID-19 cases, hospitalizations, and deaths. The Netherlands has accurate and easily accessible data that allows for a high-resolution and precise estimation of the relationship between weather conditions and the spread of viral diseases. As time trends of viral transmission can vary substantially in time and at sub-national level, the combination of high spatial and temporal resolution captures the heterogeneity of viral spreading across time and space.

In our study, parametric and non-parametric correlations are calculated for specific humidity and influenza/SARS-CoV-2 at country-level. An inverse (negative) relationship is observed between specific humidity and the incidence of influenza between 2015 and 2019, but a positive correlation is found between specific humidity and the reported cases of COVID-19. Since the positive correlation between specific humidity and COVID-19 may be caused by the use of aggregated data at national level—besides the lack of herd immunity—a Bayesian spatio-temporal disease model for the Netherlands is estimated at municipality and province level. This model allows to quantify the relation of COVID-19 cases with the specific humidity in the Netherlands at regional sub-national levels and over time.

The results of the spatio-temporal disease model indicate that the increase of specific humidity during the outbreak of the SARS-CoV-2 helped to reduce the risk of regional COVID-19 cases in the Netherlands. Specifically, an increase of specific humidity of one gram of water vapor per kilogram of air (1 g/kg) is related to a reduction of approximately 5% in the risk of COVID-19 cases.

## 2. Data

Weekly data of influenza reports were collected for the years 2015 to 2019 from the <u>Nivel Primary Care Database</u> (Nivel Zorgregistraties Eerste Lijn). Daily COVID-19 data (Reported Infections, Hospitalizations and Mortality) was provided by The Dutch National Institute for Public Health and the Environment (<u>RIVM</u>). RIVM is the government agency focused as Centre for Infectious Disease Control (CDC), under supervision of the Dutch Government. The COVID-19 data covers the twelve provinces and the 355 municipalities in the Netherlands, from March 13^th^ to July 9^th^.

Specific humidity (*q*) was calculated with the data from 34 weather stations, spread across The Netherlands. This data was provided by The Royal Netherlands Meteorological Institute (<u>KNMI</u>), the Dutch national weather service. Since KNMI only reports relative humidity (*h*), specific humidity *q* was calculated using the Rotronic formula^1^, which is based on the UK National Physical Laboratory guide for the measurement of humidity [19]:

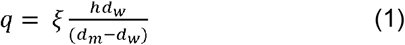

In (1), *q* is specific humidity of air vapor mixture (expressed in g/Kg), *h* is relative humidity (in percentage), *d_w_* is the density of water vapor (kg/m^3^), *d_m_* is density of the moist or humid air (kg/m^3^) and ξ = .622. Specific humidity is analyzed instead of relative humidity because relative humidity is affected by the surrounding temperature. Specific humidity in contrast remains the same in indoor and outdoor conditions^2^. The hourly weather data provided by the KNMI was aggregated at daily and weekly levels using the average values during the time periods.

Additional population data was used in the spatial-time models to calculate the incidence of SARS-CoV-2 at intra-regional level. This data was provided by the national statistical office, Statistics Netherlands (CBS) and is also used by the RIVM.

## 3. Methods

Parametric and non-parametric correlation coefficients were calculated between specific humidity *q* and the incidence of influenza and SARS-CoV-2. Spatio-temporal disease models were estimated with the daily data of specific humidity (*q_it_*) and SARS-CoV-2 (*y_it_*) in the Netherlands. Spatial dependence in the latent component of the disease mapping is modelled by specifying neighborhood relationships among the area-level risks [20-21].

In the space-time analysis, the Besag-York-Mollié (BYM) spatio-temporal model [22] is used to estimate the impact of specific humidity on the risk of SARS-CoV-2 (COVID-19) cases at province and municipality level. In the BYM model, the number of SARS-CoV-2 cases follows a Poisson stochastic process for each area *i* and time points *t*, 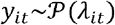, as in [23]. The parameter *λ_it_* is defined by the ecological regression:

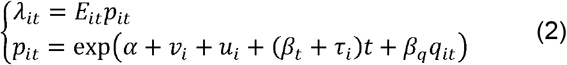

where *E_it_* is the expected number of cases in each area, at each point in time, *p_it_* is the ratio between the number of observed cases *y_it_* and the number of expected cases, *α* is the average rate of cases in all the areas, *v_i_* is the unstructured area-specific effect, which follows a Gaussian prior 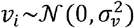, and *u_i_* is the spatially structured area-specific effect, modelled with an intrinsic conditional autoregressive prior that takes into account the number of *j*-areas which share boundaries with the *i*-areas (*j ≠ i*), i.e. the neighbors of each province/municipality [24].

In (2), the temporal component *t* is modelled with a parametric approach [25], where *β_t_* represents the global time effect, and 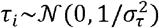 is the differential trend that captures the interaction between time and space. Specific humidity levels (*q_it_*) were included as a risk factor in the spatio-temporal model (1) to evaluate their impact on the risk of SARS-CoV-2 cases, measured by the parameter *β_q_*. When the incidence of SARS-CoV-2 per 100,000 population is used as the dependent variable, a Gaussian likelihood is used instead of the Poisson model in equation (2)^3^.

## 4. Results

Figure 1 shows the weekly historical patterns of specific humidity *q* and the incidence of influenza in the Netherlands, from 2015 to 2019. Specific humidity qduring the winter periods is between 3 and 6 g/kg, while in the summer specific humidity is above 8 g/kg. The highest incidence of influenza infection is observed in weeks with a specific humidity of less than 6 g/kg. Particularly, a higher incidence is observed in children of less than 4 years and individuals with 65 years or more (Figure 2a). Above 6 g/kg, a significant drop in infections is observed. The opposite is also true: the rate of influenza cases is 5.91 higher when specific humidity levels are approximately 2 g/kg compared to the average cases at 8 g/kg (Table 1).

**Figure 1.**
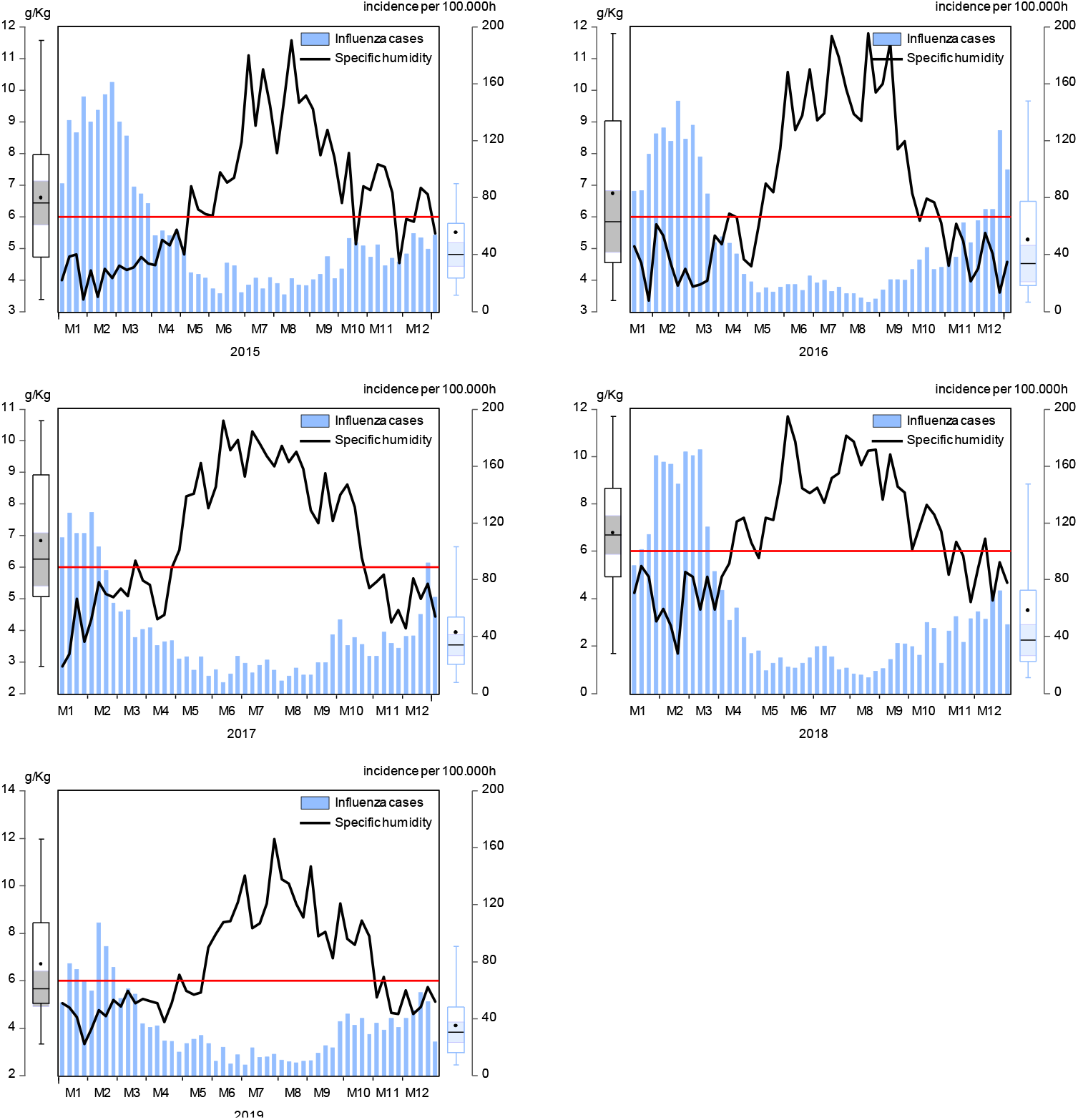
Specific humidity and incidence of influenza in the Netherlands Weekly historical observations per year.

**Table 1.** Average incidence of influenza cases at different intervals of specific humidity.

| **Specific humidity** | **Average incidence of influenza per 100000 people** |
| --- | --- |
| Below 2 g/kg | 147.3 |
| Between 2 and 3 | 135.4 |
| Between 3 and 4 | 110.9 |
| Between 4 and 5 | 79.7 |
| Between 5 and 6 | 54.4 |
| Between 6 and 7 | 33.2 |
| Between 7 and 8 | 29.5 |
| Between 8 and 9 | 24.9 |
| Between 9 and 10 | 17.2 |
| Above 10 | 15.8 |

Figure 2b illustrates the inverse non-linear relationship between the levels of specific humidity and the weekly incidence of influenza. The ordinary Pearson correlation of specific humidity with influenza cases is negative and equal to –.6986 (t-value: –15.713, p-value: 0.0000). The value of the non-parametric Spearman correlation is equal to –0.8083 (t-statistic: – 22.0993, p-value: 0.0000).

**Figure 2.**
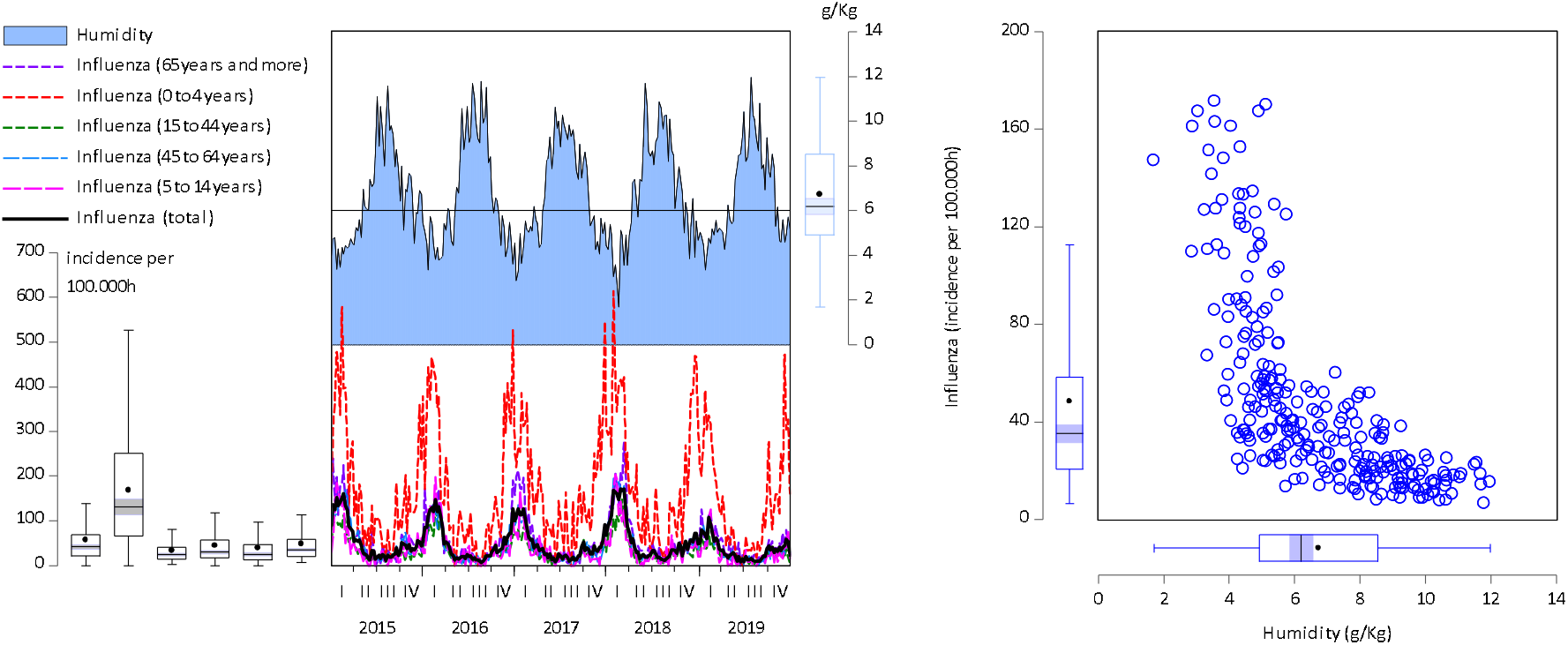
Specific humidity and incidence of influenza for age categories in the Netherlands. a: Weekly historical observations (left), b: bivariate scatterplot (right)

In the case of SARS-CoV-2, there is a positive correlation between humidity levels and the daily COVID-19 cases in the Netherlands (Figure 3). The Pearson correlation is equal to .7016 (t-value: 10.651, p-value: 0.0000), and the Spearman correlation is 0.8389 (t-statistic: 16.6734, p-value: 0.0000). This positive correlation can be explained by the lack of herd immunity—due to the novelty of the SARS-CoV-2—, the increasing number of tests, and the use of aggregate data at country level which does not capture the dissimilarities of specific humidity at sub-national levels in the Netherlands.

**Figure 3.**
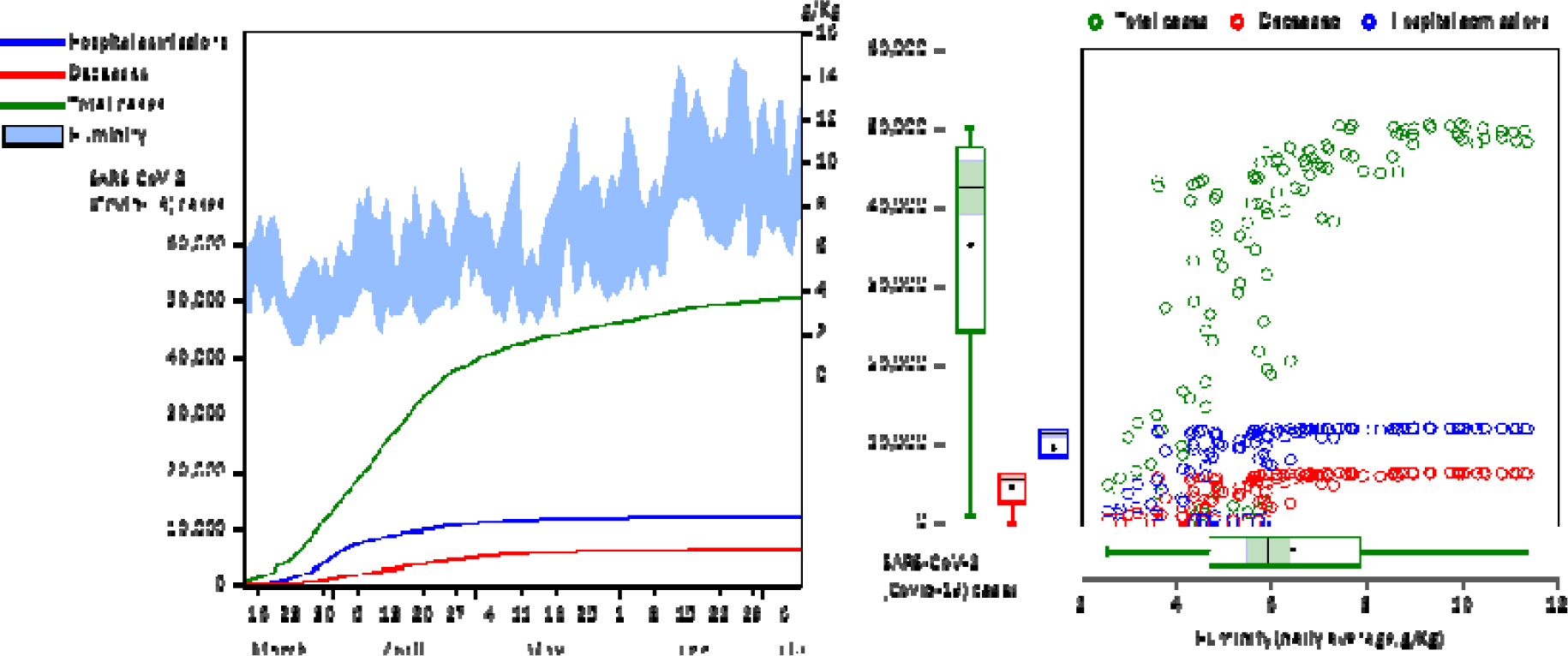
Specific humidity and SARS-CoV-2 (Covid-19) cases in the Netherlands a: Daily historical observations (left), b: bivariate scatterplot (right)

In order to take into account the differential spatial and temporal patterns of specific humidity across the Netherlands during the outbreak of the COVID-19, a Bayesian spatial-time model based on a Poisson stochastics process was estimated for the number of reported cases of SARS-CoV-2, at municipality and province level. The increase in testing, which increases reported COVID-19 cases, was controlled by relativizing the number of COVID-19 cases with the population at municipality and province level, i.e. estimating an additional Gaussian space-time model for the incidence of COVID-19 cases per 100,000 people^4^.

Table 2 shows the results of estimating the fixed-effects {*α*, *β_t_*, *β_q_* } of the spatio-temporal model. The low standard deviation of the posterior means in all the models indicates a good level of accuracy in the integrated nested Laplace approximation of the approximate Bayesian inference. The value of the exponentiated estimate of *β_t_* implies a 1.5% daily rate of infection of SARS-CoV-2 in the Netherlands from March to July of 2020, with a 95% credibility interval ranging from 1.47% to 1.62%. The negative sign of the mean estimate of *β_q_* —as well as the sign of the low and upper limit of the 95% credible interval—suggests that regions with higher specific humidity levels have, on average, lower number of COVID-19 cases in the Netherlands. Specifically, the estimated values of 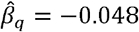 at province level and 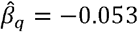 at municipality level in the Poisson space-time model indicate that an increase of one gram of water vapor per kilogram of air (1 g/kg) is related to a reduction of around 5% in the risk of Covid-19 cases.

**Table 2.** Estimation results: Besag-York-Mollie spatio-temporal disease model of daily ( ) COVID-19 cases and specific humidity ( ) in the Netherlands.

| Parameter | Area's break | Mean estimate | Standard deviation | Credible interval |  |
| --- | --- | --- | --- | --- | --- |
|  |  |  |  | 2.5% | 97.5% |
| Number of cases of SARS-CoV-2 (COVID-19) | Province | 6.770375 | 0.092920 | 6.585284 | 6.955255 |
|  | Municipality | 3.407532 | 0.043961 | 3.321127 | 3.493834 |
|  | Province | 0.015331 | 0.000354 | 0.014625 | 0.016037 |
|  | Municipality | 0.015307 | 0.000035 | 0.015238 | 0.015375 |
|  | Province | -0.047696 | 0.000361 | -0.048404 | -0.046988 |
|  | Municipality | -0.053141 | 0.000364 | -0.053855 | -0.052427 |
| Incidence of cases of SARS-CoV-2 (COVID-19) per 100,000h | Province | 67.440261 | 2.982071 | 61.584535 | 73.291041 |
|  | Municipality | 87.629212 | 3.374872 | 81.003201 | 94.249693 |
|  | Province | 2.448832 | 0.049035 | 2.352544 | 2.545038 |
|  | Municipality | 2.866350 | 0.013162 | 2.840509 | 2.892170 |
|  | Province | -6.231110 | 0.706851 | -7.619118 | -4.844281 |
|  | Municipality | -8.399374 | 0.194467 | -8.781178 | -8.017889 |

Similar results are obtained using a Gaussian space-time model for the incidence of SARS-CoV-2 cases per 100,000 population in the Netherlands. On average, regions with higher levels of specific humidity showed a lower incidence of COVID-19 cases per 100,000 people, on a magnitude of at province level and at municipality level.

Figure 4 shows the area-specific relative risks of COVID-19 in the Netherlands—at province and municipality level—calculated with the random-effects estimated with the spatio-temporal models. The regions located in the North of the country, where specific humidity tends to be higher, shohow a lower risk of COVID-19 cases. In contrast the re egions with less specific humidity, in the South East o of the country, display higher levels of relative risk of SA SARS-CoV-2 cases.

**Figure 4.**
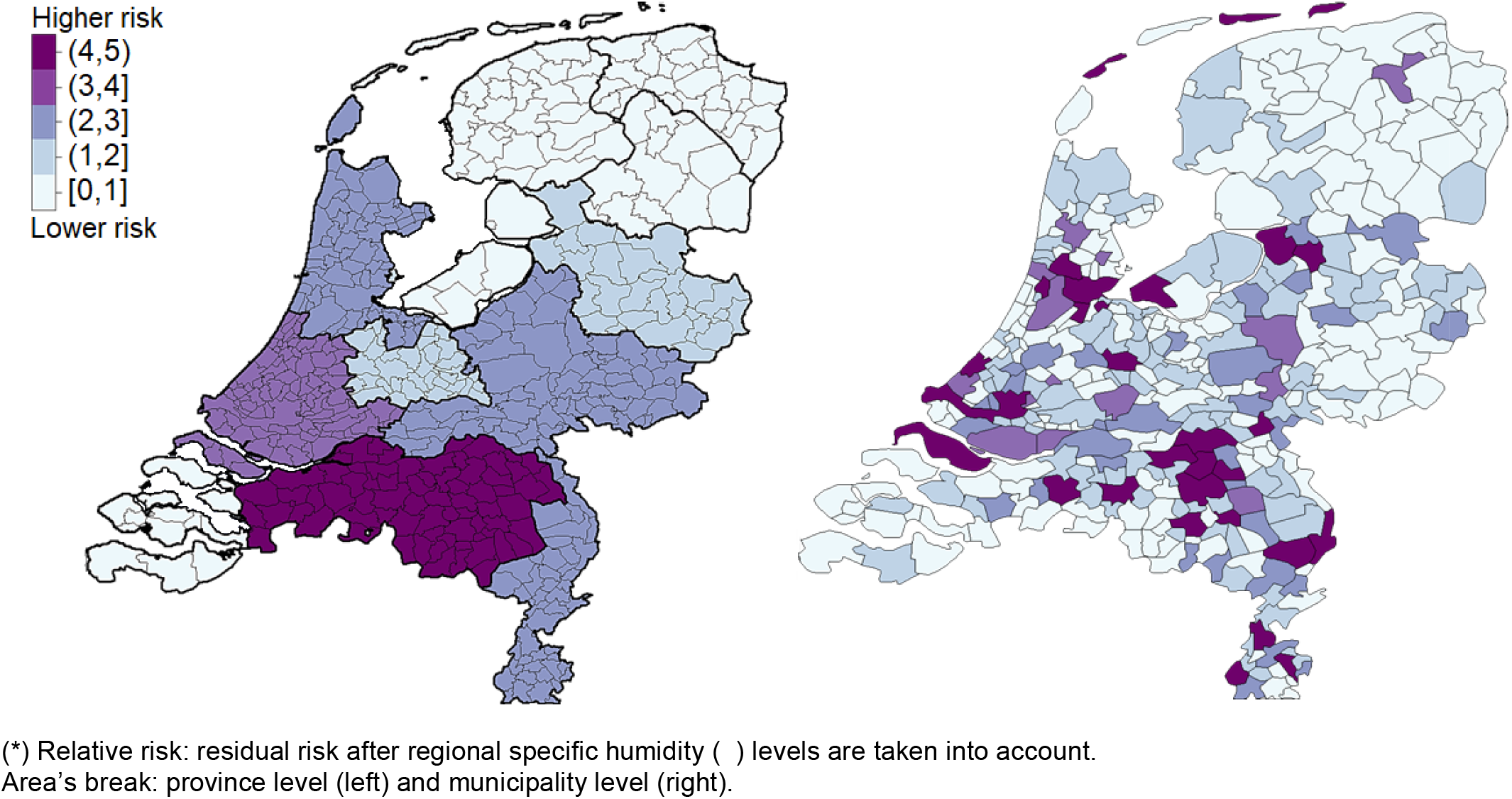
Area-specific relative risk* of SARS-CoV-2 in the Netherlands.

Interestingly, municipalities with the hiighest concentration of conservative orthodox CalalvinistProtestants in the country, i.e. those in the ‘Bible belt’, have a higher risk of SARS-CoV-2, compared to o other regions in the Netherlands. The identification of this s type of regions with a high risk of COVID-19 cases canan help to allocate resources during the SARS-CoV-2 pandndemic [26].

## 5. Discussion

Higher levels of specific humidity ( ) were found to be related to a lower number of reported cases of respiratory viruses in the Netherlands. An inverse non-linear relationship between specific humidity and influenza is observed historically between 2015 and 2019. A similar inverse (negative) relationship between specific humidity and SARS-CoV-2 cases is found using spatio-temporal models that consider the differential patterns of specific humidity ( ) and SARS-CoV-2 cases at municipality and province levels in the Netherlands. Given the seasonal dynamics of humidity in the last 5 years, an uptake (second wave) of SARS-CoV-2 infections can be expected during the weeks 43 to 45 of 2020, if low levels of humidity—below 6 g/Kg—are observed during those periods.

The results are in line with previous studies [7-18]^5^, as our risk analysis shows that higher levels of humidity may result in a slower spread of COVID-19 [10, 12]. The findings are consistent with the hypothesis that suggests that SARS-CoV-2 spreads through airborne aerosolization [28], since lower levels of specific humidity lead to a higher rate of infections due to the effects of water activity on virus survivability, ssurface inactivation, salt toxicity, or changes in pH inducuced by evaporation that modify the surface glycoproteteins of enveloped viruses and subsequently compromisee their infectivity [29]. Lower levels of humidity can partiicularly lead to higher aerosol concentration when a ddry air increases the evaporation of respiratory dropletts, i.e. when droplets smaller in diameter than aa few micrometers—referred to as ‘droplet nuclei’—evapaporate to about half their initial size [30]. In the cacase of influenza, for example, the probabilities of exposurure and infection risk of aerosol and droplet transmissioon are within the same order of magnitude, but intranasal inoculation leads to about 20 times lower infectiviity that when the virus is delivered in an inhalable aerosol [31].

In practice, the findings suggest that public policiees that promote higher levels of specific humidification—above 6 g/Kg—can lead to significant reductions in the sspread of respiratory viruses, such as influenza and SARSS-CoV-2. For example, the deployment of hygrometers aimmed at measuring specific humidity can guide the public behavior in the face of a warning of an increased risk of infection^6^. The results also imply that the use e of air conditioning (which decreases the specific humidity in a cooled area), must be complemented with humidification during the fall and winter periods, particularly when specific humidity *q* decreases below 6 g/kg.

Future studies can study the interaction effects of sunlight with humidity, as sunlight radiation may act as and additional environmental factor in the reduction and prevention of the risk of SARS-CoV-2 [32], since solar radiation plays an important role in vitamin D production, which seems to play a role in reducing COVID-19 infections [33-34].

## Data Availability

The data and codes to replicate the study are available upon request

https://www.researchgate.net/publication/343726333_Data_and_R_code_to_replicate_the_spatio-temporal_model_of_SARS-CoV-2_in_the_study_Environmental_risk_factors_of_airborne_viral_transmission_Humidity_Influenza_and_SARS-CoV-2_in_the_Netherlands_Ravelli

## Acknowledgements

The authors are grateful to Drs. Maurice de Hond, social geographer with a specialization in statistics, and Pierre Capel, professor emeritus in experimental immunology at the Utrecht University in the Netherlands, for providing us with valuable research papers and feedback about our study. Rami Alkhatib, assistant professor of mechatronics at Rafik Hariri University, did a parallel analysis on the same dataset and got similar results, supporting our findings. The data used in the study was obtained with the help of staff members at RIVM, KNMI, CBS, Rotronic, and Mariëtte Hooiveld, J Hendriksen and Korevaar JC from Nivel data (www.nivel.nl/surveillance).

## Footnotes

1 The Rotronic technical note about humidity definitions is freely available at: 0https://www.rotronic.com/media/productattachments/files/h/u/humidity_definitions_weba.pdf?_ga=2.192612136.342104324.1597340506-398495933.1597340506

2 For example, weather stations may report a temperature of 5°C and a relative humidity of 35% for the outside environment (equal to a specific humidity of 1.87g/kg). However, inside the households, the temperature can be 21°C with a relative humidity of 12% due to heating systems, but specific humidity will remains the same (1.87g/kg) regardless of the temperature.

3 The data an R codes to replicate the results with R-INLA (www.r-inla.org) are feely available upon request.

4 The Netherlands increased COVID-19 testing from 55,000 weekly tests in March to 110,000 tests per week in July 2020. The testing was only focused on care workers in March, but now the entire population is subject to testing. Due to the possible bias caused by testing, space-time models were also estimated using data of the number of hospitalizations, and similar results were obtained as those for reported cases. The results for hospitalization are available upon request.

5 Compared to these previous studies, we use more detailed data, with a high temporal and spatial resolution, in order to reduce the possibility of the ecological fallacy that may arise in the analysis of large spatial units that neglect the strong localized patterns of SARS-CoV-2 [27].

6 For example, smart social distancing regulations may take into humidity account local humidity levels, per municipality, and allow social gatherings in outside space as e.g. terraces) on humid days. At the same, social gatherings on cloudy and cool days with low humidity or in dry environments (such as restaurant), may be discouraged, unless these environments have approved humidification solution implemented. If increased humidification is not possible, air purification with HEPA filters and ionization can be alternative solutions to deactivate viral aerosols.

